# Detection and Quantification of Infectious Severe Acute Respiratory Coronavirus-2 in Diverse Clinical and Environmental Samples from Infected Patients: Evidence to Support Respiratory Droplet, and Direct and Indirect Contact as Significant Modes of Transmission

**DOI:** 10.1101/2021.07.08.21259744

**Authors:** Yi-Chan Lin, Rebecca J. Malott, Linda Ward, Linet Kiplagat, Kanti Pabbaraju, Kara Gill, Byron M. Berenger, Jia Hu, Kevin Fonseca, Ryan Noyce, Thomas Louie, David H. Evans, John M. Conly

**Affiliations:** University of Alberta, Edmonton, AB, Canada; University of Calgary, Calgary, AB, Canada; Alberta Health Services, Calgary, AB, Canada; Alberta Public Health Laboratory, Alberta Precision Laboratories, Calgary, AB, Canada

**Keywords:** SARS-CoV-2, viability, titer, COVID-19, Vero cells, environment, transmission, sputum, droplets, minimum infectious dose

## Abstract

Few studies have assessed for infectious SARS-CoV-2 in multiple types of clinical and environmental samples. In almost 500 samples from 75 hospitalized and community cases, we detected infectious virus with quantitative burdens varying from 5.0 plaque-forming units/mL (PFU/mL) up to 1.0×10^6^ PFU/mL in clinical specimens and up to 1.3×10^6^ PFU/mL on fomites including facial tissues, nasal prongs, call bells/cell phones, dentures, and sputum deposits with confirmation by plaque morphology, PCR, immunohistochemistry, and sequencing. Expectorated sputum samples had the highest percentage of positive samples and virus titers (71%, 2.9×10^2^ to 5.2×10^5^ PFU/mL), followed by saliva (58%, 10 to 4.6×10^4^ PFU/mL), and cough samples without sputum (19%, 5 to 1.9×10^3^ PFU/mL). We also detected infectious SARS-CoV-2 from patients’ hands (28%, 60 to 2.3×10^2^ PFU/mL) but no infectious virus was found in continuous speech samples despite finding high levels of infectious virus in the associated nasopharynx, throat, or saliva specimens. We demonstrated infectious virus stability in clinical samples, including those dried for prolonged periods of time. Infectious virus correlated with time since symptom onset with no detection after 7-8 days in immunocompetent hosts and with N-gene based C_t_ values ≤ 25 significantly predictive of yielding plaques in culture. One PFU was associated with ∼10^5^ copies of N gene RNA across a diversity of samples and times from symptom onset. Clinical salivary isolates caused illness in a hamster model with a minimum infectious dose of ≤14 PFU/mL. Our findings of high quantitative burdens of infectious virus, stability even with drying, and a very low minimal infectious dose suggest multiple modes of transmission are exploited by SARS-CoV-2, including direct contact, large respiratory droplet, and fomite transmission and in the context of a high binding avidity to human cellular receptors, offer an explanation of the high contagiousness of this virus.

**Research in Context:** *Evidence before this study:* We searched the literature for articles that reported on the presence of infectious SARS-CoV-2 in patients’ samples from clinical and environmental sources. We found several key primary studies and systematic reviews providing valuable background on the carriage of infectious virus and the correlation with cycle threshold (C_t_) and/or RNA copies/mL on PCR testing. Clinical correlations with respect to underlying clinical conditions and details on the onset of illness were not commonly reported with respect to the timing of obtaining specimens for culture. Few studies carefully assessed the presence of infectious virus in cough samples, sputum, nasal secretions, hands, and common high touch surfaces. A few published works were found on factors which may be associated with shedding of infectious virus.

*Added value of this study:* We assessed the presence of infectious virus shedding in almost 500 specimens from 75 patients with COVID-19 in both the hospital and community setting. High titers of infectious virus were detected in multiple clinical and environmental samples. The longest duration of recovery of infectious virus in a fomite sample was from a dried facial tissue found at a patient’s bedside table, used at least 9 hours earlier. Cough specimens revealed infectious virus in 28% of specimens with infectious virus titers as high as 5.2×10^5^ PFU/mL. Hand samples contained infectious virus with titers ranging from 55 to 2.3×10^2^ PFU/mL. Infectious viral loads correlated with N-gene based C_t_ values and showed that C_t_ values ≤ 25 were predictive of yielding plaques in culture. These experiments also showed that infectious virus is most often recovered during a 7 to 8-day period following illness onset in immunocompetent persons, and during that time the ratio of RNA/PFU in these clinical specimens varies relatively little, with a ratio ∼160,000:1. Infectious virus may be recovered for weeks to several months in immunosuppressed persons. We also showed that virus recovered from saliva specimens, representing a commonly encountered fomite sample, caused infection in the Syrian hamster model, hence demonstrating the infectiousness of the virus sourced from this type of specimen. A challenge dose as low as 14 PFU/mL yielded infection in this model.

*Implications of all the available evidence:* We have shown that SARS-CoV-2 is relatively easy to culture when obtained early in the course of illness and there are high levels of cultivatable SARS-CoV-2 in multiple types of clinical specimens and common fomites, including high-touch surfaces and demonstrated their infectiousness in a mammalian host. Our results demonstrate the presence of high quantitative burdens of SARS-CoV-2 in sputum, saliva, and droplets from coughing, which would lend support to large respiratory droplet transmission, hands which would support direct contact transmission, and fomites which would promote indirect contact transmission. We were unable to detect any infectious virus in continuous speech samples which suggests that brief conversations, without coughing or sneezing, pose little risk of transmitting SARS-CoV-2. Our findings provide an explanation for the high contagiousness of this virus and support current public health measures and infection prevention and control guidelines including physical distancing, hand hygiene, masking, and cleaning and disinfection.

## INTRODUCTION

Since February 2020, severe acute respiratory syndrome coronavirus-2 (SARS-CoV-2) has gripped the globe (Dhama et al. 2020). In response, public health and social measures were implemented based on the best available data related to the presumed modes of transmission of this virus and based on recommendations for other respiratory viruses in community and hospital settings (Centers for Disease Control and Prevention 2020; World Health Organization 2020b, 2014).

The modes for SARS-CoV-2 transmission are considered to occur through multiple routes including large respiratory droplets, contact (direct and indirect i.e. fomites), and small particle aerosols, with close contact being a major risk associated with transmission (World Health Organization 2020c). There has been debate about the degree to which respiratory secretions of varying particle sizes, including those produced by exhaled breath, may be responsible for transmission of the virus (World Health Organization 2020a; Ma et al. 2021; Klompas, Baker, and Rhee 2020; Greenhalgh et al. 2021) in part due to confusion over the relationships between a PCR signal and how that result relates to the underlying quantities of viral non-genomic RNA, virus genomes, and infectious virions. The transmission of SARS-CoV-2 is further clouded by uncertainty over the minimal infectious dose in humans although classical human volunteer studies with the 229E coronavirus have shown clinically evident attack rates as high as 50% with extremely low inoculation doses of 0.6-1.5 TCID_50_ (Reed 1984; Bradburne, Bynoe, and Tyrrell 1967).

Recent reports suggest that there is little evidence to support transmission of SARS-CoV-2 through contaminated surfaces (Lewis 2021; Goldman 2020) and the United States Centers for Disease Control and Prevention updated its guidance in May 2021 to state that surfaces are not a significant mode of transmission of SARS-CoV-2 (Centers for Disease Control and Prevention 2021). However, extensive surface contamination with SARS-CoV-2 by a symptomatic patient has been demonstrated in a hospital setting (Ong et al. 2020). In this study, a link was established between the presence of environmental contamination and the quantity of SARS-CoV-2 RNA, or cycle threshold (C_t_), detected in the clinical sample, and day post-symptom onset and shedding of infectious SARS-CoV-2. Additional studies investigating the kinetics of shedding of infectious virus from COVID-19 patients consistently report that infectious virus shedding is highest early in the course of infection (Wolfel et al. 2020; van Kampen et al. 2021; Lescure et al. 2020; Bullard et al. 2020). Given this latter observation regarding infectious SARS-CoV-2 shedding, it appears likely that patients early in the course of COVID-19 could more readily transmit and contaminate surfaces in the clinical and community setting, leading to an increased risk of virus transmission (Jefferson et al. 2020). Our study was driven by the hypothesis that COVID-19 patients in the early stage of their illness would shed infectious SARS-CoV-2 in respiratory secretions and contaminate surfaces that can contribute to transmission of the virus. We conducted detailed virological assessments of infectious virus loads in COVID-19 patients at different stages of disease, assessing various clinical and environmental (fomite) samples taken from the hospital and community setting, to gain a further understanding of the potential modes of SARS-CoV-2 transmission and to explore the reasons why this virus is so contagious.

## METHODS

### Participants and Setting

Patients who were admitted to one of four Calgary, Alberta (AB), Canada hospitals (Foothills Medical Centre, South Health Campus, Peter Lougheed Centre, and Rockyview General) and one Edmonton, AB hospital (Misericordia Community Hospital) between April 22, 2020, and March 31, 2021, and with a COVID-19 diagnosis confirmed by reverse transcriptase real time-polymerase chain reaction (RT-PCR), were approached to participate in the study. For community participants, the responsible Medical Officer of Health Alberta Health Services (AHS) Public Health (PH) in Calgary provided a list of people who had tested positive for COVID-19 confirmed by RT-PCR and were approached by telephone to participate in the study in their community setting. All participants provided informed consent for the use of any previous clinical samples completed for COVID-19 testing, collection of additional clinical or environmental samples, and other clinical information for research purposes. The study was approved by the University of Calgary Conjoint Research Ethics Board (REB20-0444). Details of the infected cases were collected by health professionals who were a part of the investigative team and included demographic background, medical history, symptom assessment, date of symptom onset, date of COVID-19 testing, and disease course. Depending on whether the individual was in the community or the hospital, the data was gathered from personal interviews or from a review of AHS inpatient records and/or PH community contact tracer reports (Epidemiologic Summary Reports). A comprehensive assessment tool covering symptoms and signs (new or worsening) associated with COVID-19 was used for all inpatient cases, including core respiratory and gastrointestinal symptoms and signs and a COVID-19 expanded list including headache, muscle/joint pain, fatigue/extreme exhaustion, nausea/sudden loss of appetite, conjunctivitis/red eye/conjunctival edema, loss of/change to sense of smell or taste and any additional COVID-19 symptoms at the clinician’s discretion (e.g. cutaneous manifestations such as “COVID toes”). This assessment was supplemented by review of medical inpatient records and assessment by an innovative electronic clinical decision support COVID-19 symptom monitoring tool administered up to three-times daily. Serial follow up was used as required for outpatients, to improve sensitivity and reduce anchoring, recall, selection and inadequate follow up bias (Meyerowitz et al. 2020). Patients were selected based on a range of early and late symptom onset and C_t_ values. Soon after the study was initiated it was recognized that patients with low C_t_ values (≤25) were more likely to produce cultivatable virus and these cases were preferentially enrolled.

### Clinical Samples and Sample Collection

Clinical specimens and environmental samples were collected and tested using RT-PCR (Berenger et al. 2021) and quantitative SARS-CoV-2 plaque assays (Harcourt et al. 2020). Clinical specimens included nasopharyngeal swabs (NP), throat swabs (TS), saliva, endotracheal aspirates (ETT), sputum if available, cough samples (spontaneous or requested), continuous speech, and clinical tissue samples and blood if appropriate. NP were collected using Flexible Mini-tip FloqSwabs (Copan) and TS using Puritan polyester or Copan ClassiqSwabs. Saliva (∼1 mL) or ETT (1 to 2 mL) were collected into sterile containers. For each of these specimens, 3 mL of Copan UTM-RT (Code 330C) or Dulbecco’s Modified Eagle’s Medium (Gibco) with addition of 2% fetal bovine serum, 1 µg/mL meropenem and 1µg/mL Amphotericin B (DMEM+) was added.

Methodologies for sample collections from the patient environment were assessed for ease and feasibility in the first few patients. Environmental samples were collected in a similar manner using an NP (Flexible Mini-tip FloqSwab, Copan), a sterile cotton-tipped throat swab, a sterile 2×2 cm cotton gauze, small pledgets cut from contaminated cloth samples (e.g. face cloth), or facial tissues to which 2 to 8 mL of DMEM+ was added. Cough and continuous speech samples were collected in sealable, transparent polyethylene bags (∼18×19 cm or 27×27 cm) which were opened wide enough to accommodate the mouth and lower face and held in place at a distance <2-4 cm to ensure adequate sample collection. Prior to closing and sealing, 2 to 5 mL of DMEM+ was added and residual air was expelled. Samples were processed in a biosafety cabinet for retrieval of the DMEM+ in preparation for transport to the Biosafety Level 3 (BSL3) containment facilities. All cough samples were accepted regardless of the vigour of the cough or if sputum was produced or not. The speech samples were continuous and directly observed by experienced collecting investigators (JC or TL) over 3 to 5 minutes and only accepted if no coughing, sneezing, or saliva contamination were observed. The kiss samples were obtained by a lip touch to the inside of the polyethylene bag and were done to simulate the double to triple kisses to the cheeks that are commonly used as a greeting in many cultures around the world. Random samplings of the polyethylene bags were cultured to ensure they were devoid of contaminating bacterial and fungal microbes that would interfere with plaque assays. Hand samples were collected by washing each hand in 10 mL of DMEM+ for ∼20 to 30 seconds in large 27×27 cm transparent, sealable polyethylene bags. Facial tissue samples (discarded post nose blowing) were collected and placed in 5 mL of DMEM+. Cell phone, call bell and nasal prong samples were collected using a 2×2 cm sterile cotton gauze which was added into 3 to 7 mL of DMEM+. Denture samples were collected by adding a few drops of saline to the denture groove, swabbing the area, and then placing the swab into 1 mL DMEM+ or by using a dry swab as noted above to which 1 mL DMEM+ was added.

### Effects of Drying on Clinical Samples of Saliva with Infectious SARS-CoV-2

Saliva samples were collected into sterile containers and 1 to 2 mL aliquots were placed into sterile polystyrene Petri dishes to study the effects of desiccation and to assess virus viability. Each sample was either neat or mixed with 6 mL DMEM+ and the latter alone was also used as a negative media control. The baseline sample was stored in a sealed container at room temperature throughout the experimental period after which DMEM+ was similarly added. The Petri dishes containing saliva samples were left open to the air in the patient care room for 2 hours to enable significant drying which was visually confirmed. The process simulated dried salivary droplets which might frequently be found in a patient’s room who was ill with an acute respiratory tract infection. The dried samples were then resuspended in 2 mL of DMEM+ and transferred to a sterile collection tube.

### Hand Transfer of Infectious SARS-CoV-2

A COVID-19 patient, with an initial C_t_ value of 14.7 and 16.9 (targets 1 and 2) on the Xpert Xpress SARS-CoV-2 test (Cepheid, Sunnydale, CA) was one day post symptom onset and had cough as one of the symptoms. The patient’s left hand was vigorously cleaned by the investigator with wet paper towels, ensuring friction was applied to all surfaces. The patient was asked to cough on their right hand and then to shake their left hand within ∼ 20 seconds. The handshake was a few seconds in duration. Hand samples were collected post handshake from each hand as described above.

### Sample Transport

All the freshly collected samples were placed on ice packs within 1-to-4 hours and then refrigerated at 4° C for up to 48 hr in a secure location before being transported at 0 to 4°C to Edmonton, AB. If the samples could not be transported immediately, they were flash-frozen in a dry-ice ethanol bath and then forwarded on dry ice. Upon arrival in Edmonton, the samples were transferred to the BSL3 containment facility and assayed for infectious virus within 24-to-48 hr of their collection. If that were not possible, a few samples were snap frozen and stored at −80°C until they could be titered.

### Cell Culture and Virus Titration

Vero (ATCC #CCL-81) and Vero E6/TMPRSS2 (JCRB cell bank 1819) were cultured with Modified Eagle’s Medium (MEM, Gibco) supplemented with 100 units/mL of penicillin, 100 µg/mL of streptomycin, 0.25 µg/mL of Amphotericin B (Gibco), and 10% fetal bovine serum (Gibco). For virus culture, 2×10^5^ cells were seeded into each well of the 12-well plates one day before titering. Ten-fold serial dilutions of the virus were plated in duplicate on Vero CCL-81 cells and cultured for 3 days at 37°C in MEM supplemented with 100 units/mL of penicillin, 100 µg/mL of streptomycin, 0.25 µg/mL of Amphotericin B (Gibco), and 1% carboxymethyl cellulose (CMC) (Sigma C4888). In a few cases, detected late in the study, some slow-growing viruses that produced small plaques on Vero CCL-81 cells were also plated on Vero E6/TMPRSS2 cells (Matsuyama et al. 2020). The cells were then fixed and stained with a solution containing 0.13% (w/v) crystal violet, 11% formaldehyde (v/v), and 5% ethanol (v/v) to permit plaque counts. All sample processing in the BSL3 laboratory was conducted with the authorization of the University of Alberta’s Human Research Ethics Board (Pro00099761) and Office of Environmental Health and Safety (RES0052249).

### Confirmation of SARS-CoV-2 from clinical specimens

A SARS-CoV-2 strain (GISAID# EPI_ISL_425177) was received from the Vaccine and Infectious Disease Organization at the University of Saskatchewan and used as a positive control and plaque reference. Like the clinical isolates described in this report, this strain formed plaques that exhibited a halo-like appearance on Vero CCL-81 cells under a CMC overlay. To further confirm the identity of a subset of virus isolates, the fixed and strained plates were de-stained with ethanol and then immunostained with a 1:500 diluted rabbit anti-SARS-CoV-2 spike antibody (Prosci Inc., cat#3525). The plaques were then visualized with a 2° goat anti-rabbit IgG antibody conjugated to horseradish peroxidase (Invitrogen, cat#G21234) and a KPL TrueBlue peroxidase substrate (SeraCare, cat#5510-0052).

### RT-PCR Assays and Viral Sequencing

E gene reverse transcriptase real-time PCR (RT-PCR) for SARS-CoV-2 was performed as described by (Pabbaraju et al. 2021). Samples were considered positive when E gene C_t_ value was <35. If the C_t_ was ≥35, amplification from the same eluate was repeated in duplicate and was considered positive if at least 2/3 results had a C_t_ <41. N1/N2 US CDC PCR (Integrated DNA Technologies; 2019-nCoV RUO Kit) was performed using a Qiagen RNA extraction kit, Promega GoTaq reverse transcription quantitative real-time PCR (RT-qPCR) reagents, and Bio-Rad CFX96. All the RT-qPCR analyses conducted using N2 primer sets were performed in parallel with control wells containing known quantities of an N-gene DNA target, which permitted a conversion of C_t_ values to molecular quantities (SQ). For the current analysis, the study relied primarily upon N2-based molecular detection and quantitation. Any other samples submitted for diagnostic testing were done using Health Canada/FDA approved tests at a laboratory accredited by the College of Physicians and Surgeons of Alberta.

Amplicon-based enrichment of SARS-CoV-2 was carried out using a QiaSeq Direct SARS-CoV-2 library preparation kit (Qiagen). Briefly, viral RNA was reverse transcribed into cDNA, followed by a high-fidelity multiplex PCR reaction using two pools of overlapping SARS-COV-2-specific 250 bp amplicons that span the length of the genome. Following the addition of unique dual indices to each sample, the libraries were sequenced using an Illumina MiSeq v2 (300 cycle) kit, generating an average of 2 million reads per sample. The quality control, generation of libraries and sequencing run were all performed at The Applied Genomics Centre in the Faculty of Medicine and Dentistry at the University of Alberta.

Bioinformatics analysis was performed with CLC Genomics Workbench v21 using the “Identify Qiaseq SARS-CoV-2 low frequency and shared variants (Illumina)” workflow. Paired-end trimmed reads were mapped to the SARS-CoV-2 genome (Genbank MN908947.3). The alignment was refined using the InDels and Structural Variants module, followed by the local realignment module. Nucleotide variants were identified by a minimum coverage of five reads and a minimum frequency of 70%. Consensus sequences were generated and submitted to the Pangolin lineage assigner (https://pangolin.cog-uk.io) to determine SARS-CoV-2 lineages.

### Statistical Analysis

Statistical analysis was performed using GraphPad Prism v9 (GraphPad Software) with additional calculations performed with Excel v16.50 (Microsoft) and appropriate tests of significance applied for continuous or dichotomous variables. In all cases, a p value <0.05 was considered significant.

### Stability of SARS-CoV-2 on Surfaces, Medical Equipment and N95 Respirators

We inoculated several types of commonly used pieces of medical equipment that are often used between patients including: stethoscope diaphragm, pulse oximeter, a bedside call bell, a keyboard, and a small personal digital device cover with clinical samples of saliva and ETT secretions known to be culture positive for SARS-CoV-2. The common touch surfaces of the equipment (e.g., diaphragm of the stethoscope, inside portion of the pulse oximeter placed on the finger) were inoculated to determine the effects of desiccation on the titer of the virus over time and to mimic bedside settings where these pieces of equipment might be exposed to saliva or sputum during the course of a patient’s COVID-19. We placed 10 µL of saliva and endotracheal secretions collected from patients at titers of 2.5×10^4^ and 1×10^6^ PFU/mL, respectively, to each piece of equipment. We allowed the inoculum to dry for periods of 30, 60 and 240 minutes to mimic actual clinical scenario environments including the time to conduct an interview and complete a physical exam by a bedside clinician and a 4-hour time interval for a check by nursing staff during a nighttime shift. The inoculated pieces were reconstituted with 400 µL of DMEM + and any recovered virus was quantified by plaque assay on Vero cells. We also inoculated 10 μL of a clinical sputum specimen with a titer of 1.2×10^5^ PFU/mL obtained from a COVID-19 patient onto coupons cut under sterile conditions from N95 respirators (Halyard Fluidshield 46727 duckbill respirator, 3M 1860 half-sphere respirator (RM), and 3M 1870+ panel respirator) (Lendvay et al. 2021). As controls (0 min), virus inoculants were applied to the surfaces of the medical instrument cut-outs and immediately those cut-out segments were placed into SF MEM to eluate viruses. The outer surface of the N95 respirator was inoculated to determine the effects of desiccation on the titer of the virus over time to mimic a bedside setting where a SARS-CoV-2 infected patient may cough or sneeze and deposit saliva or sputum on the outer surface of a mask. We dried the coupons for 30 min and then eluted and quantified the virus by plaque assay as noted above.

### Transmission and Virulence Studies in a Syrian Hamster Model

Eight-week-old Syrian male hamsters (*Mesocricetus auratus*) were purchased from Charles River Laboratories, Montreal, Quebec. Hamsters are highly susceptible to SARS-CoV-2 (Rosenke et al. 2020) and all of the studies were conducted in BSL3 containment with the approval of the University of Alberta’s Animal Care and Use Committee under authorization AUP00001847. The studies used two different lineages (B.1.279 and B.1.128), isolated from COVID-19 patient saliva samples, plaque purified three times and expanded once by passage on Vero CCL-81 cells. The hamsters were anesthetized with isofluorane, infected intranasally with 100 µL of SARS-CoV-2 (50 µL/nare) and returned to their cages. The animals were subsequently weighed daily as well as being swabbed on days 1, 3, and 6 within the nose and on the mouth and tongue with a polyester swab (Puritan 25-3318-U). The swabs were placed in 600 µL of Iscove’s modified Dulbecco’s medium containing 2% fetal calf serum and stored frozen at −80°. The hamsters were euthanized on day 14 and the four smaller lobes of the lungs homogenized in 2 mL of MEM using a GentleMacs M tube, followed by centrifugation for 10 min at 3000×g and 2 min at 8000×g. The supernatants were aliquoted with a portion reserved for RNA extraction and another for virus titration.

Virus culture and titration was conducted as described above. For PCR quantification, 140 µL of virus-containing sample was first mixed with 0.56 mL of “AVL” viral lysis buffer and carrier RNA and processed to extract the RNA using a QIAamp viral RNA minikit (Qiagen). Five microliters of RNA extracted from the oral-nasal swabs, or 5 µL of 1:10 diluted RNA from the lung homogenates, were then analyzed using a Promega Go-Taq One-step RT-qPCR kit and CDC nucleocapsid primer set and cycling protocol (IDT).

## RESULTS

### Patient Characteristics and Symptoms

Details of the infected cases, including basic demographic background, medical history, types of symptoms, date of onset of symptoms, date of diagnostic test, and course in the community or the hospital are provided in **Table 1**. All (41/41; 100%) of the inpatient cases were found to have symptoms and/or signs consistent with COVID-19 using comprehensive data gathering and attention to reducing any potential biases. One inpatient may be considered asymptomatic and arguably may have had no symptoms if one were to ignore the increased need for “throat clearing” but it occurred in the setting of a known exposure event to someone with COVID-19. Of the community cases, the vast majority (32/34; 94.1 %) were also found to exhibit symptoms and signs compatible with COVID-19. Of the total of 75 persons with COVID-19, 86.7% (65) had one or more “core respiratory” symptoms/signs including cough, sore throat, dysphagia, nasal congestion or rhinorrhea, dyspnea, difficulty breathing at some point during their illness. The remainder had “other” non-respiratory symptoms (e.g. fever, nausea, emesis, fatigue, fever, muscle aching, dys[a]geusia and /or dys[an]osmia) which have been associated with COVID-19. We identified only 2 persons (2.7%) with complete absence of symptoms in our dataset.

**Table 1:** Clinical characteristics of COVID-19 cases (n=75)

| **Basic Demographics** | **N (%)** |
| --- | --- |
| Age (years), mean +/- SD (range) | 53.3 +/-10.6 (1day-90 years) |
| Female | 41 (54.7) |
| Direct admission | 31 (41.9) |
| Admitted from care facility | 6 (8.1) |
| Admitted from transition housing | 4 (5.4) |
| Community (not admitted) | 34 (45.3) |
| **Co-Morbidities\*** |  |
| Cardiac disease and/or hypertension | 27 |
| Metabolic disorders (other than diabetes) | 27 |
| Musculoskeletal diseases | 18 |
| Respiratory | 15 |
| Mental health | 11 |
| Diabetes (Types 1 & 2) | 13 |
| Malignancy | 13 |
| Digestive disorder | 13 |
| Obesity | 10 |
| Skin and soft tissue disorders | 11 |
| Infections (non-respiratory) | 8 |
| Renal disease | 6 |
| Peripheral vascular disease | 5 |
| Solid organ transplant (< 3 months post transplant) | 2 |
| Alcohol/substance use disorder | 6 |
| No underlying comorbidities† | 10 |
| Other | 4 |

\* *Co-morbidities not mutually exclusive*
† *Mostly community participants*

| **Presenting symptoms and signs\*** |  | **N (%)** |
| --- | --- | --- |
| Symptomatic |  | 73 (97.3) |
| Asymptomatic |  | 2 (2.7) |
|  | Fever/sweating/chills | 31 (41.3) |
|  | Shortness of breath/difficulty breathing | 29 (38.7) |
|  | Fatigue/exhaustion | 33 (44.0) |
|  | Runny nose/nasal congestion | 15 (20.0) |
|  | Sore throat/painful swallowing | 27 (36.0) |
|  | Cough (productive or non-productive) | 38 (50.6) |
|  | Altered mental status | 8 (10.7) |
|  | Loss of/change to sense of smell or taste | 15 (20.0) |
|  | Muscle or joint ache or pain | 28 (37.3) |
|  | Chest pain /crackles/congestion | 7 (9.3) |
|  | Headache | 16 (21.3) |
|  | Emesis/nausea/loss of appetite | 8 (10.7) |

\* All symptoms and signs (mean of 3.5 per patient)

The symptom complexes had a variable presentation, with new symptoms/signs developing over the course of the illness while others settled but 88% of those with identified symptoms and/or signs (64/73) had at least three identified symptoms/signs. We identified 3 presymptomatic persons in our dataset where symptoms and/or signs followed within 24 to 72 hours after identification of a positive PCR test.

Core respiratory symptoms (any one of or a combination of cough, sore throat, nasal congestion/rhinorrhea, and dyspnea) were found at some point in the illness course in 85.1 % of our cohort and almost 90% had at least 3 identified symptoms/signs compatible with an expanded list of COVID-19 compatible symptoms and signs.

### Detection of infectious SARS-CoV-2 in cell culture from COVID-19 patients and their environment is associated with day post-symptom onset

A diagnostically rich sampling strategy was used to collect multiple clinical sample types and associated environmental samples from COVID-19 patients at different times post-symptom onset. SARS-CoV-2 causes a distinct cytopathogenic effect (CPE), growing quickly in culture and producing large, haloed plaques in the Vero CCL-81 cell monolayer in 2-3 days (**Figure 1**). When plated using CMC as a stabilizer, most of the strains formed large plaques that exhibited a characteristic halo structure. Variable sized plaques could be observed from different patients which were attributed to different mutations in the viruses (**Figure 1A**). A few specimens exhibited atypical plaques and on close inspection these were produced either by other viruses (e.g., Herpes simplex virus) or were artifacts caused by the destruction of the monolayer by bacteria or fungi (**Figure 1B**). For example, a specimen collected from an immunosuppressed patient with oral candidiasis, had to be passed through a 0.2 µm filter and cultured with additional anti-mycotic drugs to titer the SARS-CoV-2. Occasionally virus with typical plaques was cultured from samples with low C_t_ values after more prolonged incubation and these samples were re-titered on Vero E6/TMPRSS2 cells to ensure accurate quantification (Matsuyama et al. 2020). In two epidemiologically-linked cases the viruses formed plaques that differed in appearance from those formed on Vero CCL-81 cells (**Figure 1A**). In such cases, immunohistochemistry with an antibody directed against the spike protein was used to confirm the identity of the isolated virus as SARS-CoV-2 (**Figure 1A**). Plaque counts varied tremendously across the many different samples, ranging from near the limits of detection (∼5 PFU/mL) to >10^6^ PFU/mL. Most of the plaques were readily identifiable as SARS-CoV-2 from simple inspection of the plaque’s appearance. Selected samples of saliva-borne SARS-CoV-2 viruses were further plaque-picked, isolated, and sequenced for use in animal studies.

**Figure 1.**
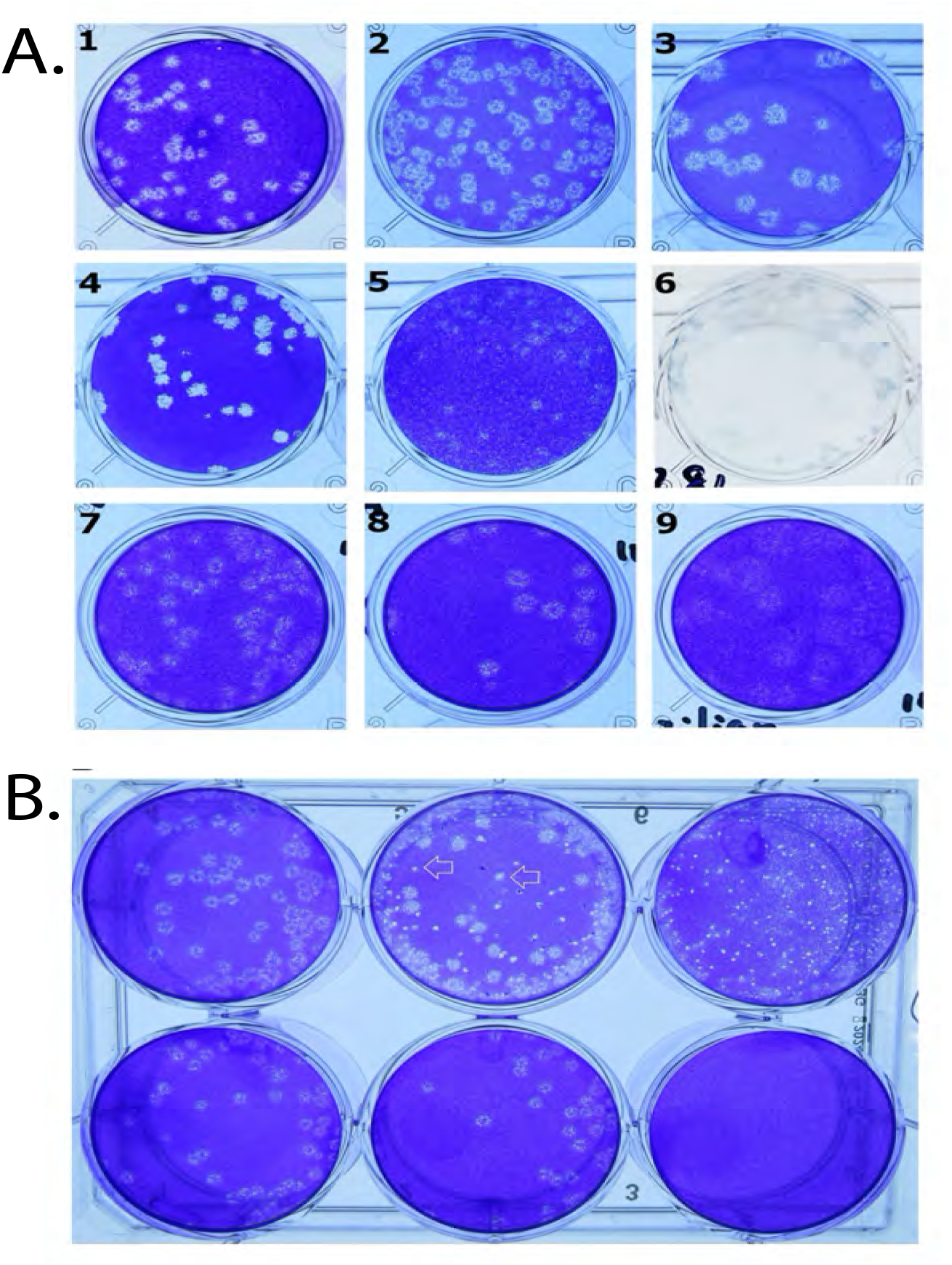
SARS-CoV-2 detection by plaque assay. <u>Panel A</u>. Dishes 1-3: SARS-CoV-2 cultured for 3 days on Vero CCL-81 cells. The virus forms different-sized plaques, but nearly all exhibit a characteristic halo structure. Dish 4: SARS-CoV-2 plaques on Vero E6/TMPRSS2 cells. Dishes 5-6: SARS-CoV-2 plaques on CCL-81 cells stained first with crystal violet (dish 5) and then destained and immunostained using an anti-SARS-CoV-2 spike antibody (dish 6). Dishes 7-9: SARS-CoV-2 variants of concern cultured on Vero CCL-81 cells (dishes 7-9, UK; South African, and Brazilian variants, respectively). <u>Panel B</u>. Cytopathic effects caused by other microbial contaminants. Patient samples were passed through 0.2 µm filters to remove bacterial and or fungal contaminants. Top row: Plaque assays performed using three different unfiltered samples. Bottom row: Plaque assays conducted after filtration. Despite incorporating broad-spectrum antibiotics and antimycotics in the culture media, arrowheads show where the growth of bacterial and/or fungal contaminants can still create plaque-like clearings in cell monolayers.

SARS-CoV-2 from each sample was also detected by RT-PCR using primer sets targeting the E and RdRP genes. A comparison of all these molecular data is shown in **Figure 2A**. There was a positive correlation between the C_t_ measured using the three SARS-CoV-2-specific probes (E versus N2, R^2^ = 0.57; RdRP versus N2, R^2^=0.46). N2 gene probes generated lower C_t_ values than E and RdRP probes. At a C_t_ of 25 with N2 primers, the corresponding C_t_ would be 1.7-2.4 C_t_ higher for E and RdRP probes, respectively.

**Figure 2.**
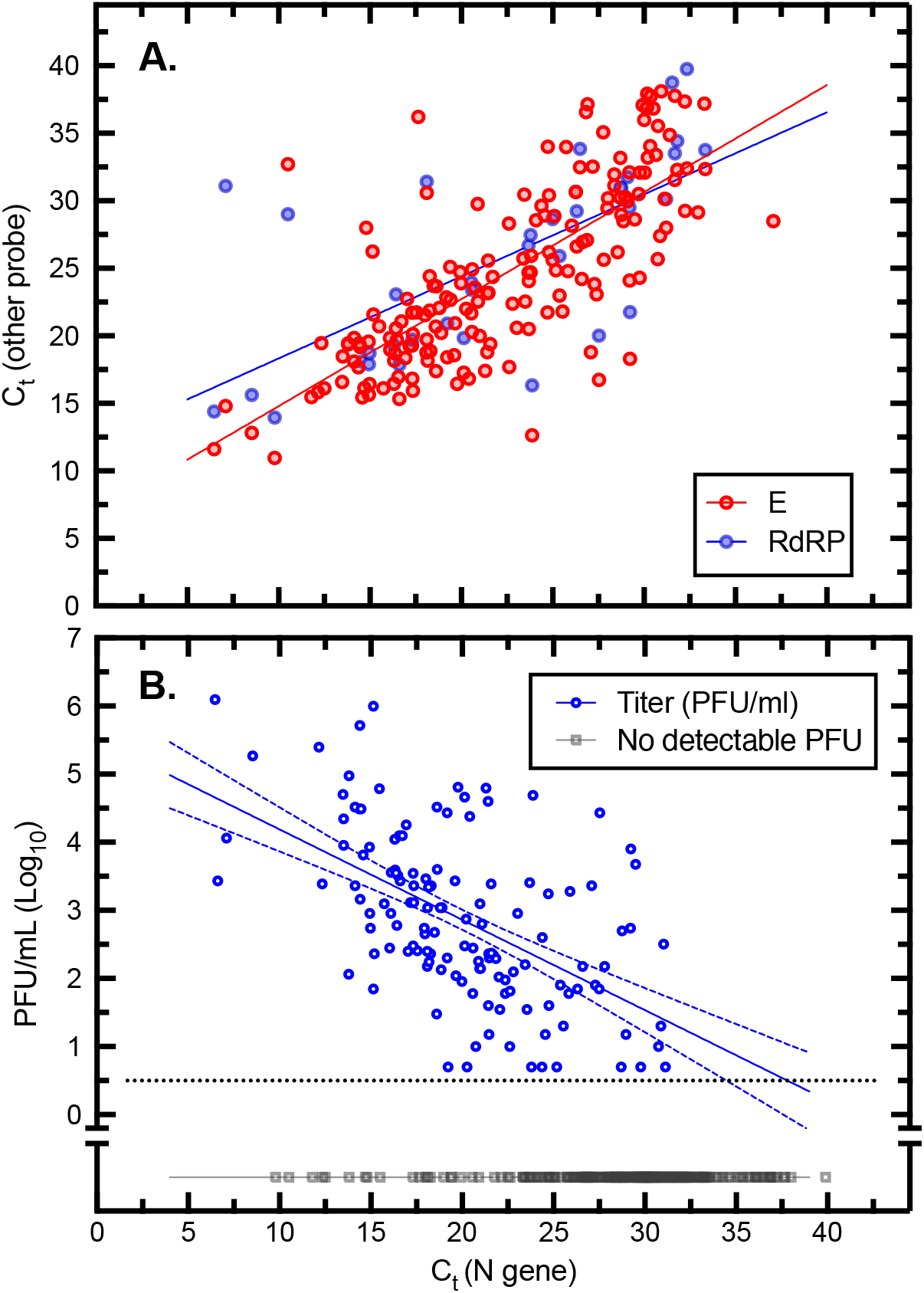
SARS-CoV-2 detection using PCR and plaque assays. <u>Panel A.</u> Comparison of reverse transcriptase qPCR assays. At the mid-point of the plot, the N gene-based assay generates C_t_ values that are about two-fold lower that C_t_ measured using E or RDRP gene primers. <u>Panel B.</u> Correlation between N gene-based C_t_ values and virus titer measured as plaque forming units (PFU). The samples exhibited a wide range of virus titers varying from >10^6^ PFU/mL to the limit of detection (∼5 PFU/mL). A least-squares fit to the log_10_-transformed data is also shown (y=-0.13x+5.5) along with the 95% confidence intervals. Specimens bearing no detectable infectious SARS-CoV-2 virus are also plotted for purposes of comparison (black squares). Most of the specimens (97%) were titered on Vero CCL-81 cells.

Overall, 33% of the plated specimens exhibited some quantity of infectious virus. A feature of the data shown in **Figure 2B** is the considerable scatter in the titers. Also clear from the plot is the potential for some clinical specimens to contain substantial amounts of virus. At the high end of the infectious spectrum, a blood-tinged sample of sputum was retrieved from a cotton gown a few minutes after being coughed up and found to have a C_t_ of 6.47 (N gene) and a titer of 1.3×10^6^ PFU/ mL. This titer is comparable to that which can be obtained when SARS-CoV-2 is cultured on Vero cells, and their related derivatives (e.g., Vero E6/TMPRSS-2 cells), where we routinely obtain titers ranging from 10^6^ to 10^7^ PFU/mL. Another specimen acquired from an endotracheal tube had a titer of 1.0×10^6^ PFU/mL and the many other specimens with titers in the 10^4^-to-10^5^ PFU/mL range show that that this level of viral load is likely common, illustrating the high quantitative burden which SARS-CoV-2 can achieve in the human respiratory tract.

We also note that the specimens reported here were obtained from patients on different dates post-symptom onset, ranging from 1 to >90 days. In this regard the date of onset was evaluated using both chart reviews and interviews. The cohort included several individuals with immunodeficiency. Except for some specimens obtained from two solid organ transplant recipients and two cases of blood-borne cancer, all the infectious specimens were detected in the first week after symptom onset (**Figure 3A**). Within a window extending up to 8-days post onset, as noted about a third of specimens contained infectious virus (**Figure 3A**). Thereafter, the C_t_ values rose to levels unlikely to yield plaques and indeed, no plaques were found. However, we continued to detect viral nucleic acids for another 2 or more weeks in some patients (**Figure 3B**). The singular exceptions to this general rule were seen in patients characterized by some type of immunodeficiency. Two of these were solid organ transplant patients, with a persistent viremia that eventually responded well to Remdesivir treatment combined with a reduction in their immunosuppressive regimens (Rajakumar et al. 2021). We also collected specimens from a patient with follicular lymphoma who had infectious virus from saliva and NP swabs which were plaque positive (1.5×10^2^ PFU/mL) 5 months from the date of original symptom onset (**Figure 3A**) with C_t_ values in the 27-28 range (**Figure 3B**). Collectively, our data illustrate the importance of timing of the sampling of clinical specimens to define the window of infectivity in immunocompetent patients and the extended window in those who are immunocompromised.

**Figure 3.**
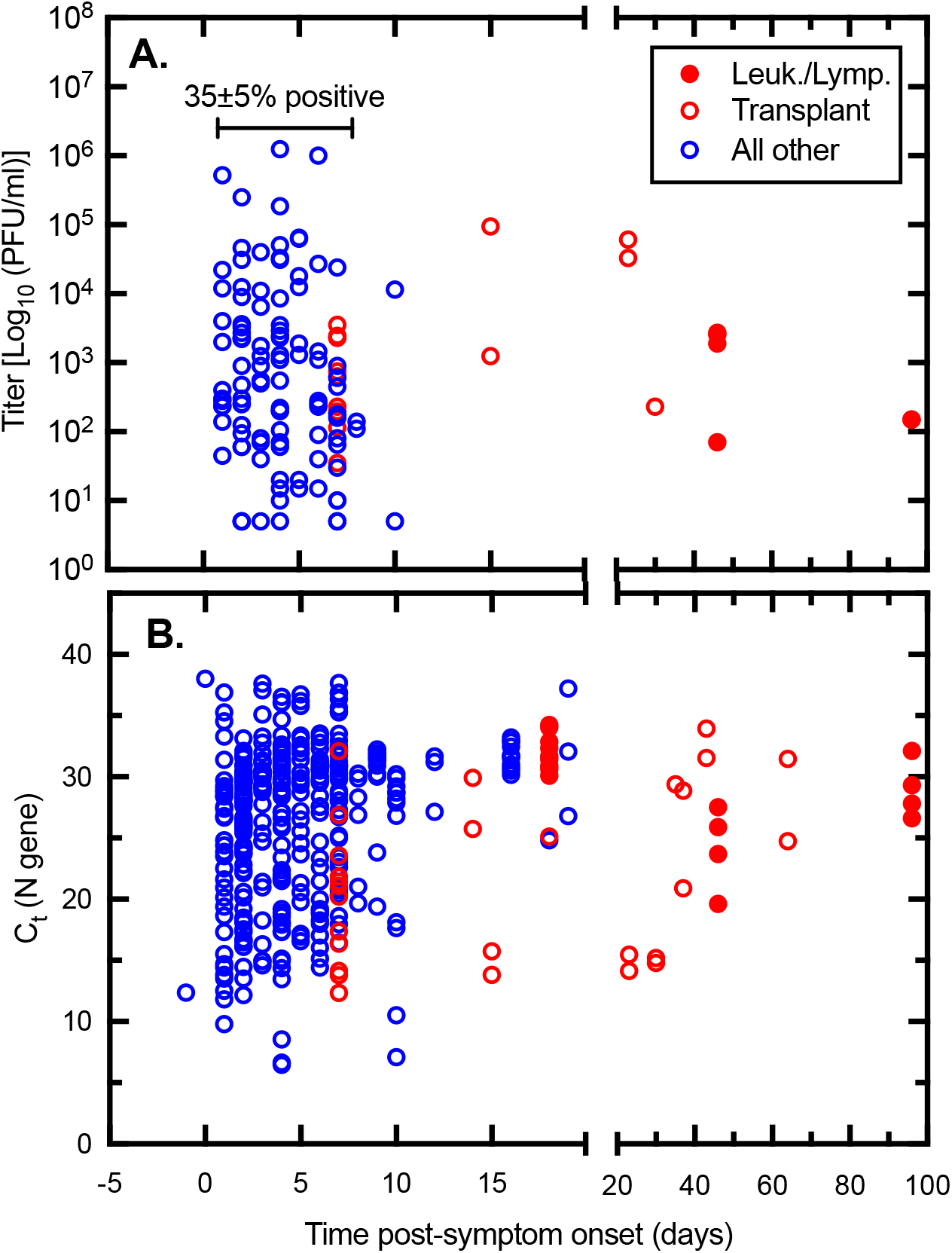
Impact of sample timing on SARS-CoV-2 virus detection. The time post-onset was calculated from interviews and/or chart review. <u>Panel A.</u> Virus titer where it could be detected. <u>Panel B.</u> All the C_t_ measurements acquired over the study. In most cases, the capacity to detect virus drops off precipitously about a week after case onset (blue data points). However, both RNA (lower panel) and PFU (upper panel) are detected for many days or weeks later where the patient is immunocompromised (red data points).

### Detection of infectious SARS-CoV-2 in clinical, environmental, and direct contact specimens

Expectorated sputum and unproductive cough samples, nasal secretions, hand samples, continuous speech samples and environmental samples collected from patients in the hospital and infected persons in the community setting were among the materials assayed for infectious SARS-CoV-2 (**Figure 4**). Of the samples from patients with concomitant NP or TS specimens positive for infectious SARS-CoV-2, the expectorated sputum samples had the highest percentage of positive samples, and highest virus titers (71%, 2.9×10^2^ to 5.2×10^5^ PFU/mL), with saliva being the next most infectious sample (58%, 10 to 4.6×10^4^ PFU/mL), followed by cough samples without discernible sputum (19%, 5 to 1.9×10^3^ PFU/mL). The presence of infectious virus with high quantitative burdens was found in 28% of all productive and non-productive cough specimens, which would have contained droplet particles of many different sizes. Some patients who did not have cough as a part of their symptom complex were asked to produce a cough which was not as natural as an illness associated cough which may have affected the positivity result. No infectious virus was found in any of 33 continuous speech samples with a known NP or throat swab positive for infectious virus, including multiple specimens with very high titers. We also detected infectious SARS-CoV-2 in samples acquired from washes of patients’ hands with culture media (28%, 60 to 2.3×10^2^ PFU/mL) and two kiss samples were found to be positive as well (11%, 35 and 70 PFU/mL). We found one tissue specimen from a placenta with infectious virus at a titer of 2.8×10^2^ PFU/mL from a mother who had had active COVID-19 infection at the time of delivery.

**Figure 4.**
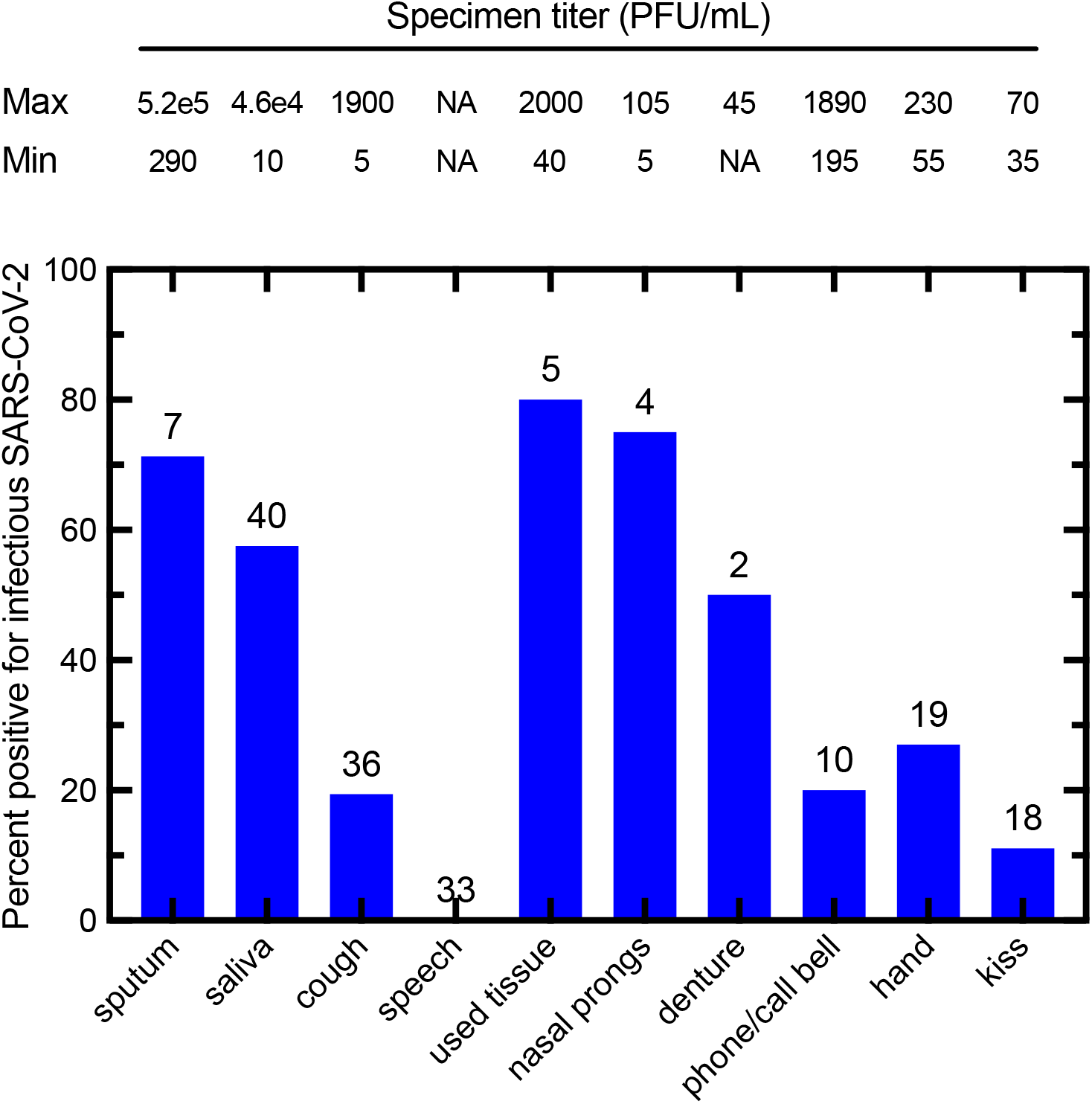
Percentage of clinical and environmental samples positive for infectious SARS-CoV-2 from patients with positive NP or TS. The number of samples acquired in each category are indicated above each bar, along with the minimum and maximum infectious titer (PFU/mL) observed for each sample along the top of the figure. Not all sample types were collected from every patient. Sputum indicates productive cough samples.

The environmental samples that were most likely to contain infectious virus were used facial tissues (80%, 40 to 2.0×10^3^ PFU/mL), followed by nasal prongs (75%, 5 to 1.1×10^2^ PFU/mL), and dentures found at the bedside which were lying there for approximately 4 hours (50%, 45 PFU/mL). Some phone and call bell samples were found to contain infectious virus (20%, 2.0×10^2^ to 1.9×10^3^ PFU/mL) although no data was collected on the frequency of cleaning of these items. Other miscellaneous environmental samples were collected over the evaluation period including a used face cloth found lying on the bedsheets with 1.2×10^2^ PFU/mL and a plastic bedrail swab which had a sputum sample deposited there by the investigator and was allowed to dry for 30 minutes with an infectious titer found of 1.5 x10^3^ PFU/mL. No infectious virus was detected on either of two pulse oximeters sampled from patient rooms.

### Hand transfer of SARS-CoV-2 reveals no decay in infectious virus titers

The COVID-19 patient who engaged in the handshake experiment was 1 day post symptom onset, had an NP swab C_t_ of 18.6 and was shedding SARS-CoV-2 as indicated by an infectious titer positive TS of 4.0×10^3^ PFU/mL and a cough sample (which contained some sputum) of 5.2×10^5^ PFU/mL. After coughing onto their right hand and then shaking the cleansed left hand, both the primary inoculation hand and the receiving hand samples were positive for infectious virus with similar findings of 1.4×10^2^ and 3.0×10^2^ PFU/mL, respectively. Although circumstances made possible only a limited single experiment, the result does illustrate the capacity to transfer the virus from hand to hand under the specified conditions.

### Effects of drying on the titer of SARS-CoV-2 in clinical samples of saliva

We examined how stable SARS-CoV-2 might be in dried saliva on items that are used in patient care settings. The saliva samples dried over 2 hours on an uncovered sterile plastic surface at room temperature (**Figure 5A**) and visually appeared to no longer to have any viscosity. After being resuspended in DMEM+, the drying step had essentially no effect on the virus titer relative to that measured in samples that had not been dried, or samples that were first diluted in DMEM+ and stored at room temperature for 2 hr. All the titers exceeded ≥10^3^ PFU/mL in the different control and experimental specimens (**Figure 5B**). As a further illustration of the stability of SARS-CoV-2 in the patient care setting in dried secretions, infectious virus was recovered from a used facial tissue that had been overlooked on a side table in a COVID-19 patient care room for 9 hr (40 PFU/mL on TMPRSS2 cells).

**Figure 5.**
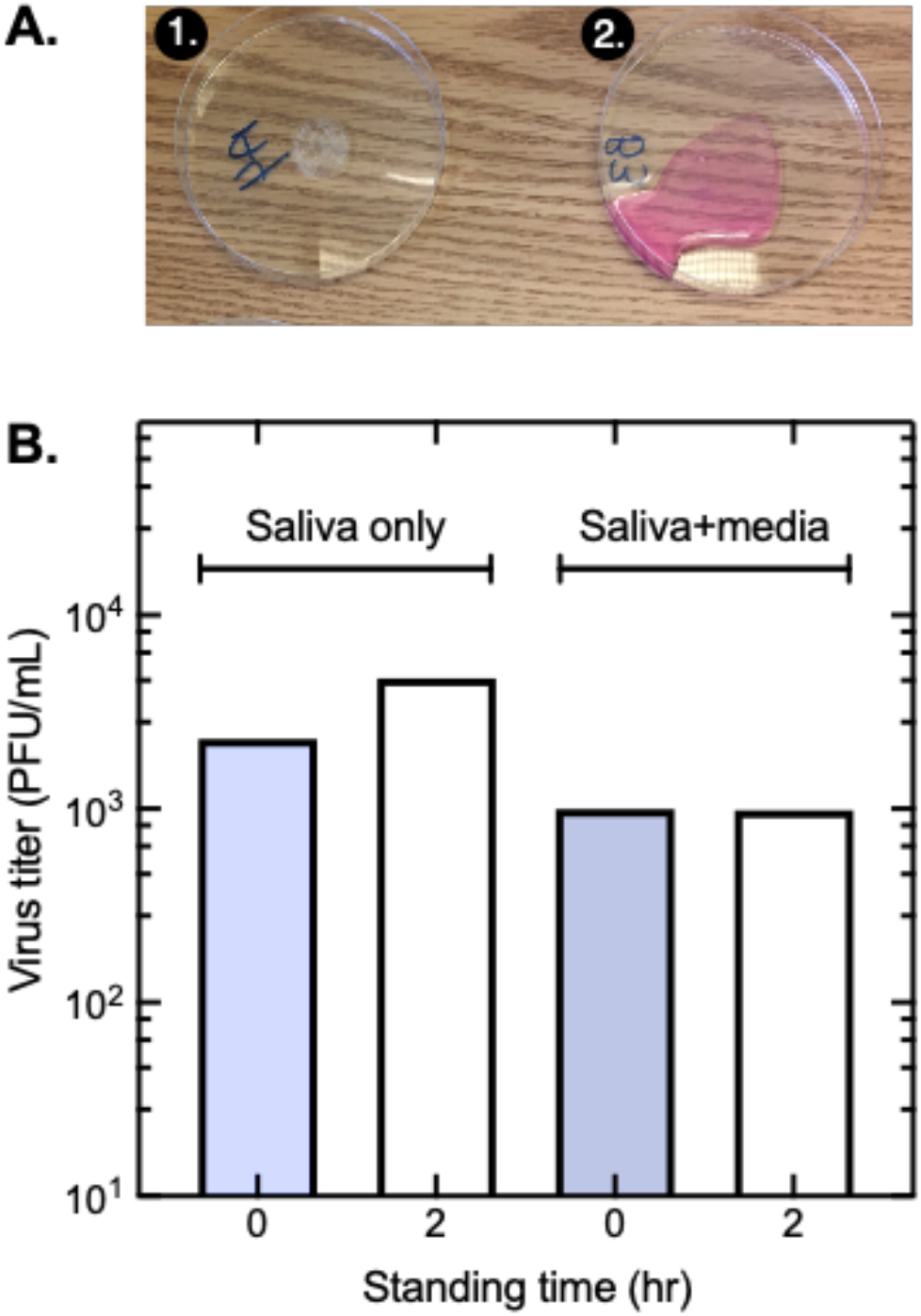
Impact of drying on infectious titer in a representative saliva sample. Saliva from a COVID-19 patient, or saliva mixed with DMEM+serum, were left in open Petri dishes in a patient care room. <u>Panel A.</u> Image showing the effects of leaving a saliva sample (dish 1), or saliva mixed with DMEM+serum (dish 2) for 2 hours. The saliva specimen dried completely. <u>Panel B.</u> Effect of standing time and drying on virus titers. The two control samples (blue bars) were stored on ice in closed tubes during the two-hour experiment.

### Virus stability on clinical equipment

There was no difference in the number of viruses eluted from the stethoscope diaphragm, pulse oximeter, bedside call bell, and keyboard at 0 min (**Figure 6**). However, the amount of virus eluted from a personal digital device cover was significantly lower than the other four instrument cut-outs (ANOVA test, p=0.0495). Presumably it is because the materials of the cover hinder recovery of the virus or there were residual virucidal chemicals present on the surface. Infectious virus was recovered from three of the commonly used medical instruments (call bell, computer keyboard, and stethoscope diaphragm) for up to 4 hours; for at least one hour on the pulse oximeter and 30 min from personal digital device cover (**Figure 6**). We also spotted 10 µL samples of sputum, each containing 1.2×10^3^ PFU of SARS-CoV-2, onto coupons cut from a N95 respirator. This was done to mimic a patient care setting where a COVID-19 patient may cough or sneeze and deposit saliva or sputum on the outer surface of a healthcare worker’s personal protective equipment. After being left to dry in room air for one hour, an average of 6.7×10^2^ PFU (55%) of the virus was recovered from each spot, demonstrating that SARS-CoV-2 retains infectivity in dried sputum on the outer surface of a N95 respirator. We further examined samples of cough droplets (i.e., macroscopically visible droplets) captured in transparent polyethylene bags, to examine the relationship between C_t_ values measured in NP swabs and cough samples (**Figure 7A**). Each datum point was further color-coded to show the viral culture quantitative burden. Patients with a high virus load (low NP C_t_ values) and early in the course of their illness (5.5±2.9 days post-onset, n=8) produced droplets with relatively high titers of infectious virus. Cough droplets and sputum arise from both the upper and lower respiratory tract, although sputum generally represents a larger, more semi-solid respiratory-type secretion and is more easily collected. Most of the sputum specimens exhibited low C_t_ values (<20) (**Figure 7A**) and 71% had infectious virus (**Figure 4 and Figure 7A**). An analysis of saliva specimens showed a similar pattern (**Figure 4 and Figure 7B**), although in contrast to the cough/sputum samples a low C_t_ in the NP specimen was not always predictive of whether virus would be detected in saliva samples **(Figure 7B)**.

**Figure 6.**
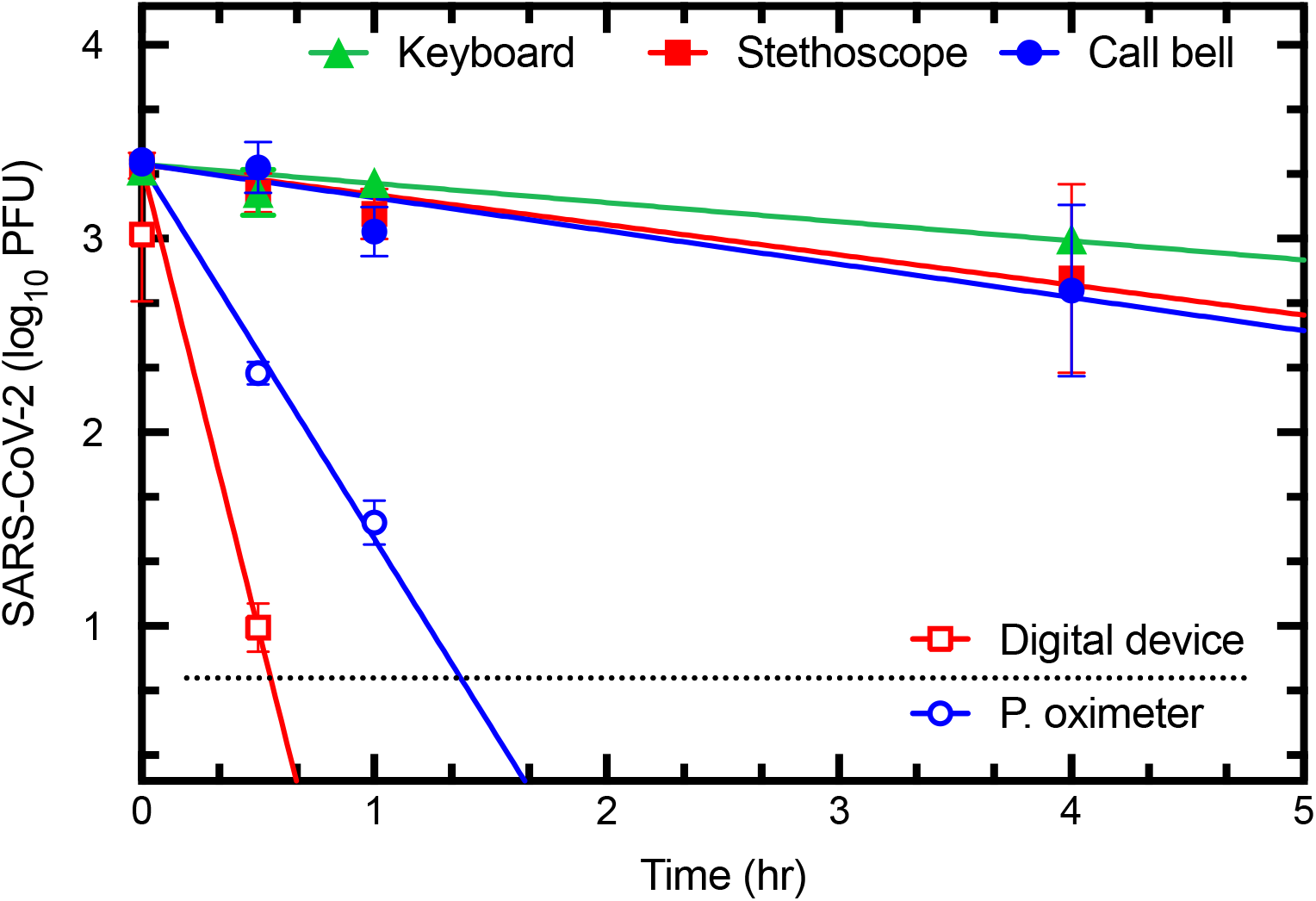
Stability of SARS-CoV-2 on representative patient-contacted surfaces. The indicated items were acquired from the wards and transferred for testing without further treatment beyond everyday maintenance and cleaning. Some were partly disassembled to facilitate safe handling and access. An endotracheal tube sample, containing 1×10^6^ PFU/mL SARS-CoV-2 diluted in DMEM, was applied in three 10 µL volumes to each item and either retrieved immediately, or stored in a biocontainment hood for the indicated times before recovery and plaque assay. The half-lives range from 3 min (digital device cover) to 82 min (keyboard).

**Figure 7.**
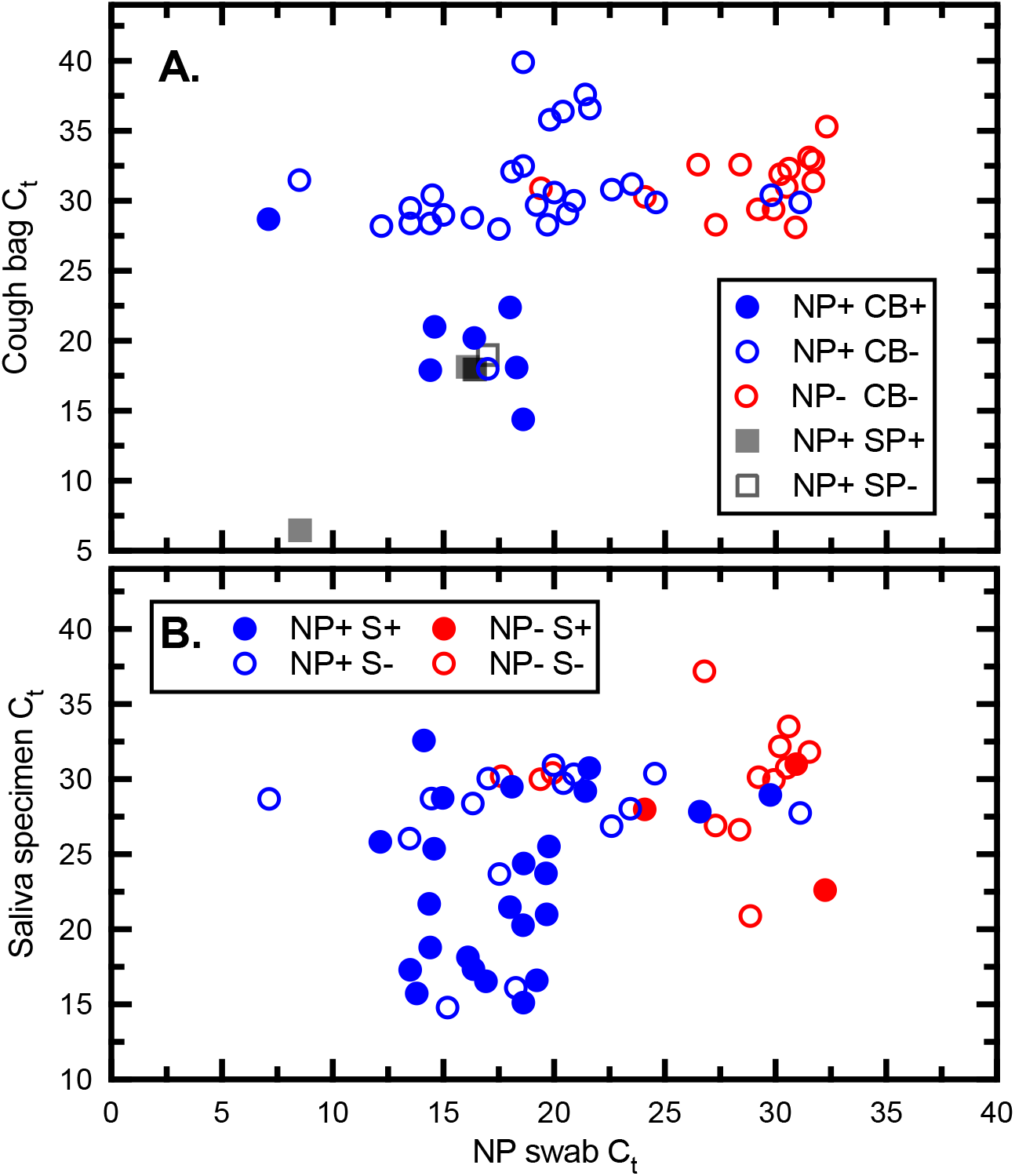
Infectious SARS-CoV-2 in saliva, sputum, and cough specimens. <u>Panel A.</u> Virus samples were acquired from NP swabs as well as cough bag samples (CB) or as sputum (SP). Samples determined to contain infectious virus are indicated with solid-colour coding. <u>Panel B.</u> Virus samples were acquired from NP swabs as well as saliva. A C_t_≤25 in the NP swab predicts that about 1/3 to 1/2 of the cough/sputum and saliva specimens will also bear infectious virus, respectively.

### Correlation of C_t_ (N gene) with Infectious Virus Titers

We also examined how well C_t_ is predictive of infectious titer (**Figure 8**). Samples that contained infectious material exhibited significantly (P<0.0001) lower C_t_ values compared with plaque negative specimens. Infectious samples exhibited a mean C_t_ of 19.2, whereas the mean in non-infectious samples was a C_t_ mean equal to 29.5. These data show that in practice one is unlikely to detect virus plaques where the samples exhibit a C_t_ >25 based on the assays used in our platforms. For the very rare exception where plaques were detected at a C_t_ > 25, it was a single plaque. Below a C_t_ of ≤25 (N gene) the odds improve significantly, where 80% of samples we studied contained infectiousvirus. This C_t_ value offers a convenient benchmark that may be useful when evaluating the potential transmission risk posed by clinical and environmental specimens.

**Figure 8.**
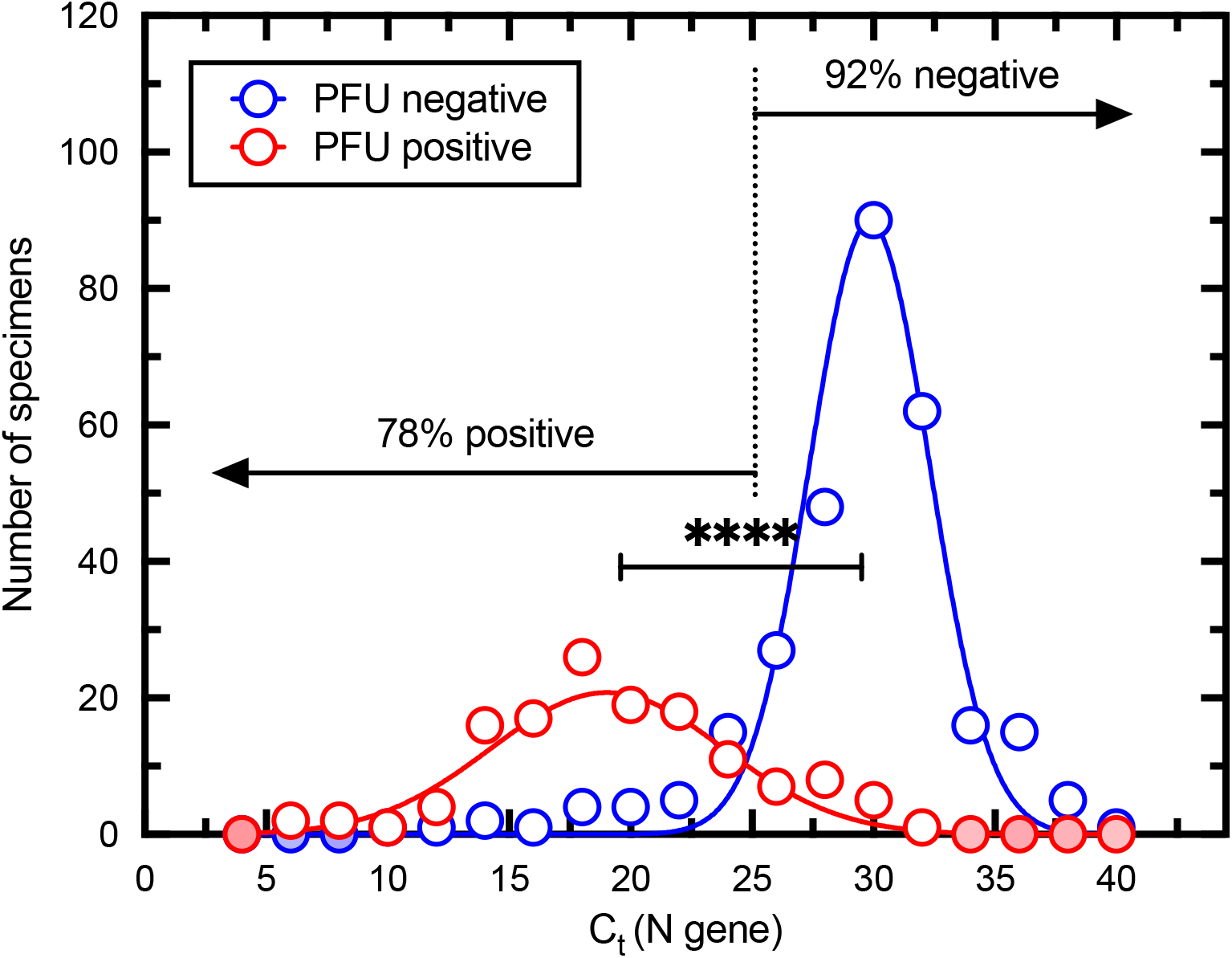
Relationship between Ct and positive plaque assays. The specimens were divided into plaque-positive, and plaque-negative categories and the distribution of Ct values calculated using bin steps of two. A non-linear fit of two Gaussian curves to these data is also shown. A sample with a Ct ≤25 is 78% likely to bear infectious material while samples with C_t_> 25 are 92% negative. An unpaired t-test, using Welch’s correction for unequal variances and sample sizes, indicates that the two means (19.0 versus 29.8) are significantly different (two-tailed P<0.0001). Red or blue filled shading indicates that the binning process counted no specimens in these bins.

### SARS-CoV-2 RNA to PFU Ratio

Another striking feature of the data, which has been previously reported by others (Plante et al. 2021), is the very high ratio of viral RNA to PFU. This can lead to misunderstandings regarding the health risk posed by samples with high C_t_ values. To calculate this parameter, all the qPCR assays incorporated additional wells containing known quantities of the N gene template. This allows one to estimate of the starting quantity (SQ) of viral target sequences by comparing C_t_ values. This data, combined with the titer and known volumes of materials assayed, permits an estimate of the RNA/PFU ratio. **Figure 9A** illustrates this point where we have calculated the number of RNA copies (from the SQ/mL data) per plaque forming unit (from the PFU/mL). We observed a Gaussian distribution of values centered on a mean of 10^5.2±0.8^ (i.e., 160,000) RNA targets per PFU. The method is not ideal as it makes assumptions about the efficiency of RNA extraction, PCR amplification, and plating. However, it isn’t greatly different from ratios calculated using the more homogeneous virus that can be harvested from culture (∼10,000:1).

**Figure 9.**
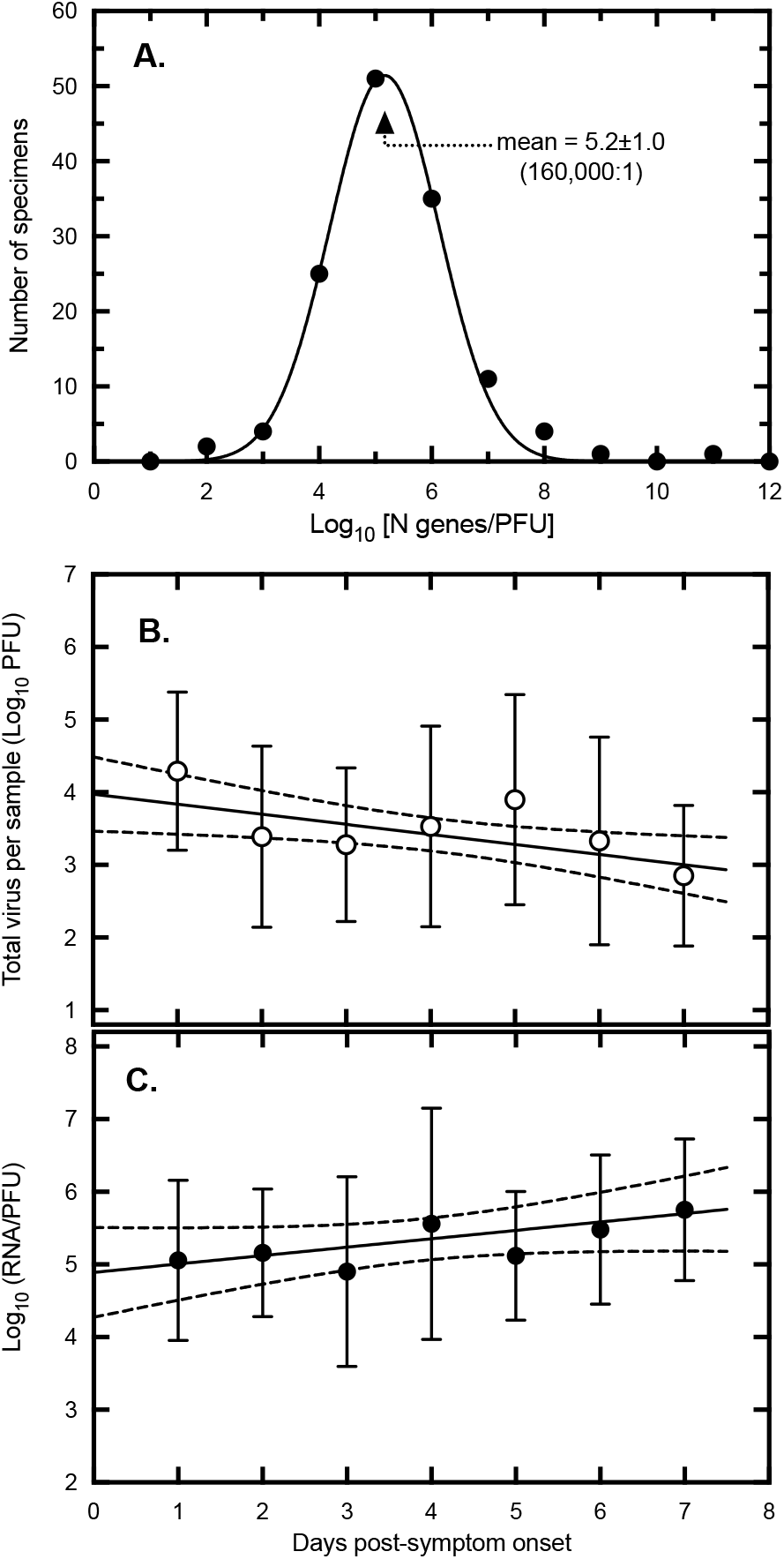
Relationship between viral RNA quantity, PFU, and sample timing. Panel A. The ratio of RNA to PFU was calculated using the virus titer plus a determination of the number of N-gene copies across all of the specimens. The ratios were log_10_ transformed and the distribution calculated across bin steps of one log_10_. A non-linear fit of a Gaussian curve to these data is also shown, centered on a mean of 5.2 ± 1.0. This represents 10^5.2^ = 160,000 RNA copies per PFU across all of the samples. <u>Panel B</u>. The total quantity of virus in each infectious specimen was calculated using the titer (PFU/mL) and the known collection volumes (i.e., specimen + carrier/diluent). These values were then averaged across all of the samples for each of the indicated days. The plot shows a least-squares fit to the log_10_-transformed data along with the 95% confidence intervals. The negative slope is significantly non-zero (p=0.015). Error bars represent standard deviation. The average virus load declined about 10-fold over the course of a week beginning at ∼10^4.0^ PFU/mL on a hypothetical day zero. <u>Panel C.</u> The relative infectivity was calculated using the virus titer (PFU/mL) and specific quantity of virus RNA/mL in each sample. The positive slope is significantly non-zero (p=0.032). Error bars represent standard deviation. The ratio of RNA/PFU increased about 8-fold over the same seven days, starting at ∼10^4.9^ RNA/PFU on day zero.

**Figure 9A** was compiled from all the infectious specimens acquired over a range of days post-infection and this led us to wonder whether the timing of sample collection might have had an impact on the RNA/PFU ratios. To do this we focused on the specimens collected in the seven-day window post-symptom onset (**Figure 3**), but still comprising a variety of specimen types. We observed that the average of the total quantity of virus detected in each of the specimens declined about 10-fold over the week after symptom onset (**Figure 9B**). At the same time the RNA/PFU ratio rose ∼8-fold (**Figure 9C**). For practical reasons it is difficult to sample on the day of symptom onset but extrapolating these plots back to a hypothetical day zero shows that the highest virus loads (PFU) and most infectious virus (RNA/PFU) might well be found at that time point.

### Virulence Studies in a Syrian Hamster Model

Syrian golden hamsters are highly susceptible to SARS-CoV-2 (Rosenke et al. 2020). To document the *in vivo* infectivity of these clinical specimens, two different virus isolates were plaque purified from saliva samples and expanded once on Vero cells. They were then used to inoculate hamsters by the intranasal route. **Figure 10** shows the results of this experiment. Both isolates were highly infectious in hamsters, with a dose as low as 14 PFU causing the transient weight loss that characterizes this model (**Figure 10A**). Nasal swabs were collected at days 1, 3, and 6 post-infection and detected virus replication that reached titers as high as 10^4^ PFU per swab, far more than the input doses. By day six post-infection, infectious virus could no longer be detected (**Figure 10B**). Nor were any plaques recovered from the lung homogenates at the end of the experiment (day 14) although some residual RNA was still detectable (not shown). Typically, the peak of infection was delayed a few days with the two lowest doses of virus, it was three days post-infection with infectious doses of 14 or 30 PFU, but just one day with the two highest doses. The levels of virus RNA paralleled the virus titers (**Figure 10C**). We also calculated the ratio of RNA to PFU in samples acquired on days 1 and 3, these ranged from 4,000 to 100,000 RNA/PFU in different animals and were perhaps ∼5-fold higher, on average, on day 1 than on day 3. Unfortunately, the small sample size precludes drawing further conclusions from the study. However, it is apparent that these low-passage SARS-CoV-2 specimens are highly infectious even at low doses and exhibit the same high ratios of RNA to PFU detected in human clinical specimens.

**Figure 10.**
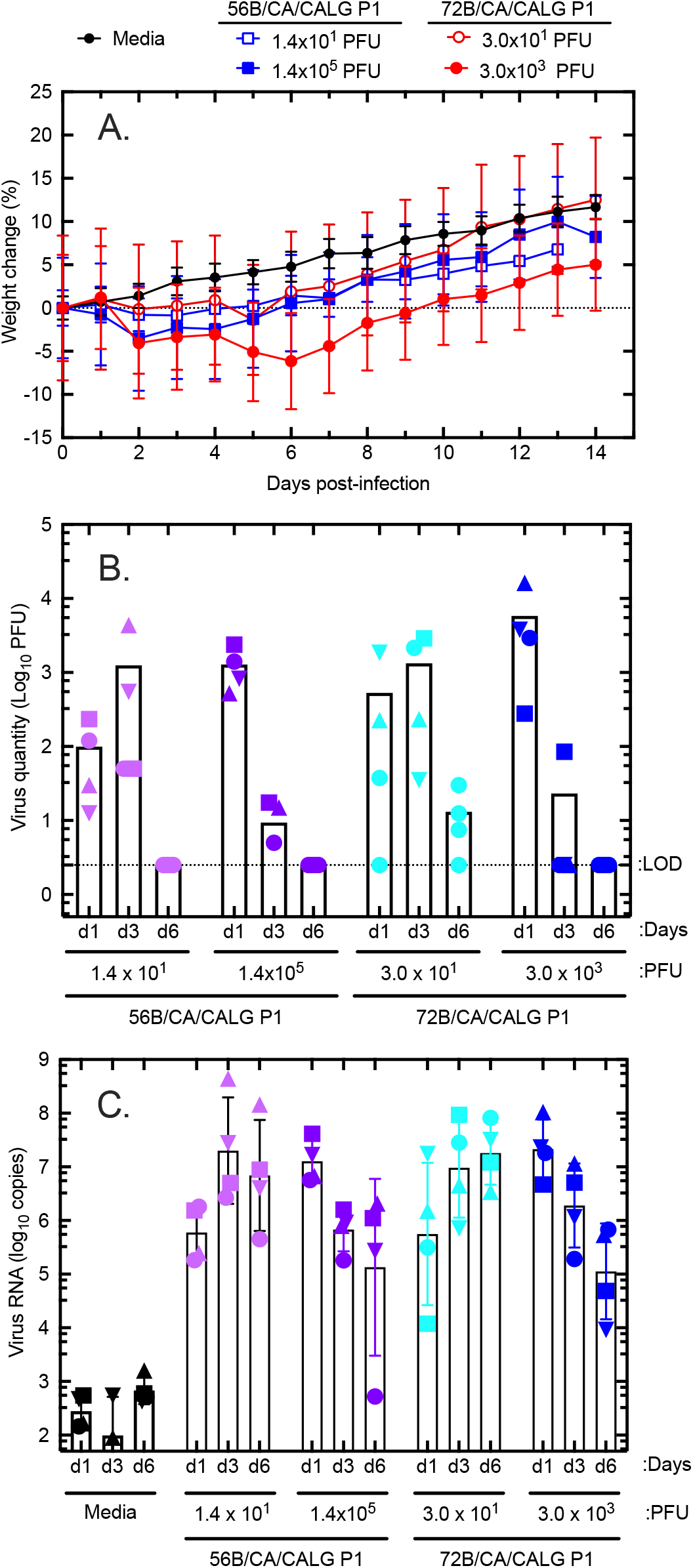
Virulence testing in Syrian hamsters. Two of the virus specimens (56B and 72B) were plaque purified and bulked up to higher titers with one passage. The two stocks were then used to inoculate four groups of hamsters (4 per group) with the indicated doses of virus. Four control animals were also inoculated by the same intranasal route, with an equal volume (100 µL total) of serum-free media. <u>Panel A</u> shows the weight change relative to the starting weight for the animals in each of the five groups. Error bars represent standard error of the mean. A nasal swab was collected from each animal on days 1, 3 and 6, post-inoculation, and assayed for virus by plaque assay and virus RNA by qPCR (<u>Panels B and C</u>, respectively) Both virus specimens produced weight loss and high titers of intranasal virus were detected at days 1 and 3 post-infection. Characteristically the lower doses (14 PFU for 56B and 30 PFU for 72B) yielded the most virus on day 3 post-infection, whereas the higher doses induced the highest levels of infection immediately after challenge, on day 1. The RNA is more persistent than virus, it could still be detected 6 days post-infection in all of the infected animals, whereas no virus could be detected at this date. The animals were euthanized at day 14. LOD = limit of detection.

## DISCUSSION

Our data demonstrated that the timing of collection of clinical specimens is of great importance relative to finding infectious virus. We found that infectious SARS-CoV-2 could be detected using Vero CCL-81 cell cultures with relative ease for an average of 4-5 days after symptom onset but that this infectivity declined dramatically 7 to 8 days after symptom onset in both immunocompetent community and hospitalized inpatients. Selecting patients early in the course of their illness with lower C_t_ values confirmed our hypothesis that this diagnostic rich sampling strategy maximizes the likelihood of detecting infectious virus and illustrates the relatively narrow window of infectivity beyond which one is unlikely to recover infectious virus. This finding is consistent with other studies which have reported that infectious virus is shed at highest levels from SARS-CoV-2 infected persons early in the course of infection (Young et al. 2020; Wolfel et al. 2020; van Kampen et al. 2021; Lee et al. 2020; Folgueira et al. 2021; Bullard et al. 2020). Our findings also illustrate the risk posed to others regarding the maximal transmissibility period by most COVID-19 patients and serve to underscore the importance of potentially prolonged transmissibility in those with immunodeficiency syndromes which has been reported in other literature (Decker et al. 2020; Baang et al. 2021).

In line with the recommendations on systematic symptom assessment and serial follow up (Meyerowitz et al. 2020), our comprehensive interviewing strategy and serial follow up found that over 97% of all patients had symptoms and/or signs although our dataset was predominantly adults with only 2 pediatric cases. There was no significant difference whether the affected persons were hospital inpatients or from the community. Interestingly in some elderly cognitively impaired persons who would not be able to provide reliable interview responses, signs such as rhinorrhea, an elevated temperature, tachypnea or sputum production were noted by experienced health care workers which added to the precision of the findings. Multiplicity of symptoms and signs was frequent, and serial follow up and secondary interviews over time were extremely useful. We identified 3 persons who had positive test results prior to symptom onset. Our findings are congruent with a recent systematic review of higher quality studies (Byambasuren et al. 2020) which used a fixed effect meta-analysis and found asymptomatic cases represented only 17% (95% CI 14% to 20%) of COVID-19 cases. A recent study of household transmission where a comprehensive capture of symptoms was done with use of a daily symptom monitoring tool, review of classic and non-classic symptoms plus initially daily RT-PCR testing, found 100% (12/12) of COVID-19 patients were symptomatic and is very consistent with our data (Lewis et al. 2021). Our results are not congruent with earlier studies suggesting as many 40-45% of patients with COVID-19 are asymptomatic (Oran and Topol 2020) but much of the early literature was based on rapid and incomplete cross-sectional studies which were likely subject to significant bias.

We found a strong correlation between the E and N gene C_t_ values and that a C_t_ ≤25 in the SARS-CoV-2 N gene assay was strongly correlated with the likelihood of finding infectious virus in both clinical and environmental samples. The presence of relatively high viral titers in the NP, TS, sputum, nasal secretions, and saliva samples provides valuable insights into the potential modes of transmission. We found infectious virus in cough samples in 28% of the patients where the “control” NP or TS or saliva was positive for infectious virus, but not one continuous “speech” sample was found to have infectious virus. All continuous speech specimen collections were monitored closely to ensure continuous speech and no coughing or sneezing occurred during the collection period. These observations may be explained by the very high ratio of viral RNA-to-PFU that characterizes diverse clinical specimens (∼160,000:1). A specimen would need to exhibit a relatively low C_t_ (≤ 25 in this study using N-gene probes) if a C_t_ was to be used as a surrogate for predicting the presence of infectious material. It was considered unlikely that any virus contained in the “air” deposited into the bag would have been expelled given the observation that there was moisture buildup noted visually within the transparent surfaces of the bag. The mean N gene C_t_ value found from the continuous speech specimens was 30.2±2.5. Many studies have placed reliance on any C_t_ value, regardless of how high it was, as evidence that infectious material is also present in collected specimens but the actual C_t_ value and its correlation with a SARS-CoV-2 *in vitro* culture specimens is important. The public health implications of this finding would suggest that a short 5 minute conversation with an infected COVID-19 patient at short distances in the absence of coughing or sneezing would be unlikely to be responsible for transmission of SARS-CoV-2 in most settings. Although we were unable to recover any virus from the samples collected within the limits of our methods, we cannot rule out the possibility that infected persons might shed infectious SARS-CoV-2 with shouting or singing (Katelaris et al. 2021).However, there have been experimental studies conducted in a confined room with susceptible subjects exposed to rhinovirus-infected human volunteers, who sang and conducted vigorous conversations for prolonged periods and revealed no transmission by these routes (D’Alessio, Dick, and Dick 1972), but it is well recognized there are many technical collection issues and difficulties associated with detecting viruses in air samples (Verreault, Moineau, and Duchaine 2008; Pan, Lednicky, and Wu 2019).

The finding of a high quantitative burden of virus in cough and sputum samples would lend support to the transmission of SARS-CoV-2 in the large droplets that are typically generated by these events. One of the authors who collected most of the specimens (JC) noted readily visible macroscopic droplets of saliva and /or respiratory secretions deposited on the sides of the transparent collection bags in almost all cases. Our findings are congruent with a previous study which found that cough and sneeze samples in infected volunteers were found to have cultivatable Coxsackievirus A type 21 and the virus was carried in these droplets in an airflow dependent manner by large floor fans in an enclosed army barracks and infectious virus was detected in large air samplers on the side opposite to inoculated volunteers (Couch et al. 1970).

We did find infectious virus in high quantities ranging up to 2.2×10^4^ PFU/mL on hands, kisses, nasal prongs, and occasionally cell phones, call bells, gowns, dentures, and a specimen of placenta. We also demonstrated the virus can be transferred from one contaminated hand to a previously cleansed hand. Our observations suggest that kisses and human hands could be important for direct contact transmission given the high frequency of habitual human behaviours such as nose, lips and eye touching (up to 15.7 times per hour) and nose-picking (up to one of every three subjects (Nicas and Best 2008; Hendley, Wenzel, and Gwaltney 1973). This would allow inoculation of a relatively high virus burden directly onto sites bearing ACE2 receptors. This mode of transmission has previously been documented in the seminal studies (Hendley, Wenzel, and Gwaltney 1973) with reported attack rates of 36.4 % in human challenge experiments, when susceptible volunteers were asked to touch their nasal or conjunctival mucosa with fingers previously contaminated with a dried drop of Type 39 rhinovirus.

SARS-CoV-2 also appears to be environmentally stable under real world circumstances in clinical specimens. We detected very little decay in virus titers using desiccated saliva acquired from a patient who was able to provide multiple replicate specimens. The virus titer changed little relative to the baseline samples when either saliva alone, or saliva mixed with serum-supplemented DMEM, was allowed to dry for slightly more than 2 hours and we demonstrated a high burden of infectious virus from actual clinical specimens inoculated on multiple pieces of commonly used medical equipment and N95 respirator masks and a used dry facial tissue found at the bedside nine hours after the patient had left the hospital room. There are multiple *in vitro* experimental studies which have shown similar survival of SARS-CoV-2 on both porous and non-porous (e.g. plastic and polymer) surfaces over time frames ranging from several hours to four or more days (van Doremalen et al. 2020; Riddell et al. 2020; Chin et al. 2020). Our data confirms the same virus stability using actual clinical specimens directly from patients and from their immediate environment in both the hospital and community settings and provides strong support for the fomite route of transmission.

An abundance of previous studies has documented transfer of infectious respiratory viruses from inanimate objects (fomites) that have been contaminated with nasal or respiratory secretions involving transfers onto the fingertips and then to the mucous membranes of the nose, mouth, and eyes. These investigations have employed an array of human challenge studies, epidemiological studies, virologic studies and intervention studies (Boone and Gerba 2007). Rhinoviruses are responsible for the majority of human cases of the “common cold” and in one such study attack rates of 50% and 56% was observed when recipients handled coffee cups and plastic tiles, respectively, that had been previously contaminated with a clinical strain of rhinovirus by infected donors (Gwaltney and Hendley 1982). Infectious rhinoviruses are also found on high-touch surfaces in home settings, providing further support for the hypothesis that contact transmission is an important mode of rhinovirus transmission (Winther et al. 2011; Gwaltney and Hendley 1978). Human CoVs (229E, OC43, NL63, and HKU1) are the second most frequent cause of the “common cold” after rhinoviruses and may be responsible for up to 30% of common colds. Where the inactivation coefficients have been calculated and compared directly it has been reported that HcoVs OC43 and 229E exhibit a stability comparable to rhinovirus and are much more stable than influenza virus (Boone and Gerba 2007). Respiratory syncytial virus (Hall, Douglas, and Geiman 1980) has been recovered from multiple fomites and infectious virus has been demonstrated to be transferred to hands from touching these contaminated surfaces. Other studies have shown that when preparations of SARS-CoV-1, MERS and other HCoV are suspended in a matrix resembling lung cell debris to mimic natural respiratory secretions, they can persist up to nine days on inanimate surfaces, further implicating this as a transmission mode (Sizun, Yu, and Talbot 2000; Otter et al. 2016; Kampf et al. 2020). A recent study on the survival of SARS-CoV-2 mixed with mucus from the upper respiratory tract revealed a survival time analysis of 10.2-12 hours on human skin which corroborates our findings (Hirose et al. 2020). Infectious MERS was also isolated from fomites in a hospital setting including bedrails, bedsheets, an anteroom table and an IV hanger (Bin et al. 2016).

Although ethical considerations render it difficult to conduct direct challenge experiments with SARS-CoV-2, future studies with highly susceptible species like Syrian hamsters might be expected to offer important insights into whether fomites can be a significant source of infection. Our findings of extensive environmental contamination and the strong supportive evidence for fomite transmission do not align with other studies and commentaries (Zhou et al. 2020; Goldman 2020; Ben-Shmuel et al. 2020) which reported no infectious SARS-CoV-2 in the environment and discounted fomites as a risk for transmission. However, unless the sampling is designed to accommodate the limited period when patients are typically infectious (up to 7-8 days post-symptom onset) and can capture sufficient material needed to detect an infectious particle (∼160,000 PCR templates, C_t_ ≤25), such studies would be highly compromised to detect any SARS-CoV-2 because of both the biology of the virus and behaviour of the adaptive human immune response to infection and the resulting disease course.

Our findings provide unique insights into understanding the contagiousness of this virus. Some of the most infectious saliva and cough specimens contained >10^5^ PFU/mL of virus, suggesting that a source droplet of 10-100 µL might have contained 10^6^ to 10^7^ PFU/mL or 1 to 10 PFU/μL. This is comparable to the best titers of virus that can be obtained through cell culture in a controlled laboratory setting. Saliva was used as a typical “fomite” seed and the fact that purified clones of SARS-CoV-2 derived from this source caused infection in the Syrian hamster model provides direct evidence of the transmissibility of the virus to another mammalian host. It thus fulfills both Koch’s postulates and the Gwaltney-Hendley postulates of viral causation (Gwaltney and Hendley 1978; Fredricks and Relman 1996). More critically we could produce disease with doses of only 14-to-30 PFU with these isolates and the minimum infectious dose in Syrian hamsters has been reported to be as low as 1 TCID_50_ with other stocks (Rosenke et al. 2020). It is quite conceivable that the minimal infectious dose in humans is in the range of 1-5 PFU which is extraordinarily low. Experiments in human challenge studies in the Common Cold Unit from the 1960s showed that as little as 10^0.6-1.5^ TCID_50_ (∼3-20 PFU) of a HCoV could cause infection with attack rates ranging from 17-67% of inoculated volunteers (Bradburne, Bynoe, and Tyrrell 1967). Thus, very few particles (Macnaughton et al. 1980) of a relatively stable virus may be capable of transmitting SARS-CoV-2 between humans, which would favour multiple routes of transmission and contribute to its relative contagiousness.

Our study has several strengths including the large number of patients acquired in a prospective manner and the ability to carefully establish the timing of symptom onset through a detailed chart and record review of the medical interviews by experienced healthcare providers. We could detect, titer and identify infectious virus with relative ease in many samples from a diverse group of patients including immunocompromised hosts. This was made possible by using a sampling-enriched strategy that focused on patients with a low Ct value (using a diagnostic PCR E gene assay) based on work by Bullard and colleagues. (Bullard et al. 2020).

We recognize our study has some limitations. Although most large-plaque forming SARS-CoV-2 strains plate equally efficiently on Vero CCL-81 cells and on TMPRSS2 transduced cell lines, we did detect a couple of specimens that were more easily detected on the latter cells. By mostly using CCL-81 cells there may have been a few other missed isolates. Whether any manner of cell culture can replicate the “plating efficiency” that is observed in the human pharynx or lung is unknown without a proper understanding of the minimal infectious dose in humans. We were unable to collect serial specimens in many cases. Our continuous speech sampling strategy was limited by the health of the patients, and we cannot rule out that more prolonged acquisition times or singing or airflow dependent droplet carriage might detect infectious virus. However, under the conditions tested, our findings suggest continuous speech for up to 5 minutes is not associated with the detection of infectious virus. We also have limited numbers of experiments on the effects of drying on the virus infectiousness and most are relegated to the hospital as opposed to the community environment. Similarly, the hand transfer experiment was not done in replicate and there remains the possibility that there was infectious virus on the cleansed hand as no cultures were obtained before the handshake to document no pre-existing virus. However, shear forces created by the friction of the cleaning and previous studies demonstrating over a 1-2 log reduction of virus with simple hand washing with water would argue against disregarding this finding which is very biologically plausible (Bellamy et al. 1993).

We have acquired one of the largest collections of infectious samples in a SARS-CoV-2 infected population and have sought to systematically quantify the actual burden of infectious virus in clinical and environmental samples, including fomites and our evidence is compelling that contact transmission is an important and overlooked mode of transmission. Our study provides a quantitative numerical framework for evaluating the risk of encountering an infectious virus particle, given the relationships between Ct, RNA copies/PFU, and days post-symptom onset. Our findings add novel and unique findings to the literature relating to the science of the transmission of this virus. With a probable very low minimal infectious dose in humans, our detailed observations and findings would support that SARS-CoV-2 exploits multiple modes of transmission, and would suggest it is important not to focus on a singular mode of transmission. A broad array or “bundle” of mitigation strategies as outlined in the currently used public health measures which includes attention to fomite control, offers the greatest degree of protection from transmission.

## Data Availability

Data will be made available upon request.

## Acknowledgements

We are thankful to the nursing staff and other hospital staff on all the units for their co-operation in the collection of specimens in these patients. We would like to thank the patients and their families for allowing specimens to be collected in the face of moderate to severe illness in both the hospital and community settings. We thank the Alberta Precision Laboratory technologists for their assistance with testing and transfer of samples. We would also like to thank Nicole Favis and Sarah Samuelson for their technical assistance.

## Author contributions

JC, DE, Y-CL conceived of this work. BB completed the ethics submission and subsequent amendments. JH and BB facilitated the identification of patients in the community for specimen collection. JC and TL collected all the clinical and environmental specimens with the assistance of LW. Y-CL and DE provided all the cell lines and expertise in the BSL3 Laboratory. KF, KP and KG completed the E gene assay. BB, KF, KP, and KG assisted with the transfer of the specimens from the hospital site to the diagnostic laboratory and then to the BSL3. RN performed all the Syrian hamster model work. BB, KF, KP, KG, LK and LW managed the clinical and laboratory data collection and data cleaning was done by LK, LW, RM and JC. All authors were provided a draft of the manuscript for comments and were provided with an opportunity to present revisions. JC, DE, Y-CL, and RM wrote the initial drafts of the manuscript and collated comments from the other authors engaged in the work and later comments from all authors.

